# Physician burnout and symptom of anxiety and depression: Burnout in Consultant Doctors in Ireland Study (BICDIS)

**DOI:** 10.1101/2022.09.29.22280503

**Authors:** Genevieve Crudden, Fabio Margiotta, Anne M. Doherty

## Abstract

**Background:** Burnout is a work related syndrome comprising three major dimensions: emotional exhaustion, depersonalisation and reduced professional efficacy. It is recognised that burnout may be associated with psychopathological sequelae, such as symptoms of anxiety and depression.

**Aims:** This study aimed to examine the relationship between burnout and symptoms of depression and anxiety in senior doctors in Ireland. We examined personality traits as a mediating factor on burnout, in relation to work related stress among the participants.

**Methods:** This study was a national cross sectional survey of consultants. Data included basic demographic data, work-related data, burnout as measured by the Maslach Burnout Inventory, personality traits, and symptoms of depression and anxiety from the Depressive Anxiety Stress Scale [DASS}.

**Results:** Results were obtained from 477 consultants, yielding a response rate 21.9%. Nearly half (42%) reported high burnout levels. The Depression and Anxiety Stress Scale revealed high levels of stress symptoms, and lower levels of anxiety symptoms. After controlling for gender and age, depressive symptoms were associated with high levels of emotional exhaustion and a higher clinical workload

**Conclusions:** The this study reported a high level of work related burnout, associated with symptoms of depression and anxiety in senior doctors. This is concerning, and strategies are required that consider the relationship between personality and burnout. Further research to evaluate strategies to ensure physician wellbeing and optimal delivery of patient care are required to address this serious problem.

## Introduction

Burnout is an occupational phenomenon related to stress, and has been described as *“a particular response to stress involving psychological withdrawal. It is a costly coping mechanism seen as the end-point of a process which begins with the stress response”*. [1] It is defined by the Word health Organisation (WHO) in ICD-11 as *“a syndrome conceptualized as resulting from chronic workplace stress that has not been successfully managed. It is characterized by three dimensions: feelings of energy depletion or exhaustion; increased mental distance from one’s job, or feelings of negativism or cynicism related to one’s job; and reduced professional efficacy”* [2]. The WHO classify it as an occupational condition and not as a medical condition.

It can have a significant effect on the health of the individual worker and can affect how they view and approach their work: *“What starts out as an important meaningful and challenging work becomes unpleasant, unfulfilling and meaningless*.*”* [3]

In the decades preceding the Covid-19 pandemic, physician burnout has become an area of increasing concern, both from an occupational medicine standpoint, and from the perspective of patient safety. First described in 1974, the construct of burnout has been developed into a measurable three-dimensional model comprising emotional exhaustion, cynicism (depersonalisation) and ineffectiveness (low personal accomplishment). [4] It is a common problem with some studies reporting rates as high as 50%, and it may have a profound effect on the lives of the physicians affected. [5]

Doctors have been identified as a group who are at increased risk of developing work-related stress. In a large study comparing US physicians with the general population, although higher educational attainment was identified as a protective factor for burnout in the general population, a medical degree conferred an increased risk independent of hours worked. [5] The authors concluded that the occurrence of physician burnout is set apart from the larger societal trend, and that there are factors specific to physicians and their work which predispose this group to poorer outcomes. Riley suggested that burnout in doctors may be the end consequence of the interplay of challenging work environment, personality factors, subjective lack of control and insufficient reward. [6] The author suggests that the “conscientious and committed personalities” working in medicine are at a loss when faced with certain environmental strains. These strains include the “demand control imbalance” identified by Karasek, [7] along with effort reward imbalance, and inadequate support in the work environment. [8] The extent of the role of personality in relation to burnout remains unclear in general, and has not been studied in the Irish context. Previous research has indicated that personality traits of neuroticism or ‘negative affectivity’ has been associated with greater stress and burnout in doctors [9]

The impact of burnout on patient care is well established. Burnout is associated with lower patient satisfaction, involvement in adverse events, and failure across domains of professionalism which affect patient care, including empathy, interpersonal communication and adherence to guidelines. [10-12] It has been suggested that burnout may be associated with issues relating to patient safety, as both burnout and patient safety relate to organisational, social and individual factors which depend on environmental resources. [13] There is also evidence that medical errors and their sequalae can in turn cause or worsen practitioner burnout and mental health problems, an effect that may be compounded by organisational factors including an organisational culture of blame, inadequate personal support and suboptimal administrative support [14, 15]. Two key systematic reviews examined the relationship between burnout and patient safety. Hall et al identified a relationship between poor patient safety outcomes such as errors, and noted a lack of prospective studies. [14] Dewa et al reported a strong relationship between burnout and problems with safety, but the relationship between burnout and patient satisfaction with care is less well defined. [15]

Burnout is associated with increased risk of health problems in doctors: a range of difficulties including cardiovascular disease and shorter life expectancy, problematic alcohol use, broken relationships, depression, and suicide. [16] While burnout is an occupational phenomenon rather than a mental illness, there are many areas of overlap with depression, and in particular the experience of hopelessness, helplessness and powerlessness are found in individuals with burnout and with depression. [2, 17] People who are severely affected by burnout are likely to also report symptoms of depressive illness: one Finnish study reported that half of those with severe burnout were also experiencing a depressive episode. [18]

This study aimed to investigate the prevalence of burnout and associated factors and their relationship with the development of psychiatric morbidity in a cohort of senior physicians working in the Irish Healthcare system. We sought to explore the impact of burnout and personality factors on the development of depressive and anxiety symptoms.

## Methods

This study utilised a cross-sectional design, to study the prevalence of burnout in a national population of senior hospital doctors (consultants) in Ireland. The methods were previously described by Margiotta et al. [19] We will summarise here for ease of reference: an anonymous survey was distributed to members of the Irish Hospital Consultants Association (IHCA) via their electronic database. The IHCA is a representative organisation for hospital consultants across Ireland with 2160 members. An email was sent to the membership including a hyperlink to the electronic survey, the study information sheet and the consent form. Participation was voluntary, and responses were completely anonymised. Ethical approval was obtained from the Clinical Research Ethics Committee of the Saolta group. Data were collected between 1st September and 30th November 2016.

Participants were asked to provide demographic details and information and details of their current working climate and lifestyle. The questions encompassed work-related characteristics and stressors. These included the person’s medical specialty, their type of contract, satisfaction with remuneration, hours worked (weekly), time spent face-to-face with patients, and any experience of medico-legal or disciplinary action.

Burnout itself was measured by the Maslach Burnout inventory General Survey (MBI-GS) which contains 3 subscales, namely, emotional exhaustion, depersonalization or cynicism, and professional efficacy. High levels of emotional exhaustion and cynicism are associated with burnout, while high levels of professional efficacy are protective. This questionnaire is validated in medical populations of clinicians internationally. Although it is not recommended that the three internal scales of MBI-GS be combined, previous studies have created a model which adds high levels of emotional exhaustion to high levels of cynicism and low professional efficacy. [20]

The Depressive Anxiety Stress Scale (DASS) is a validated self-report instrument which measures symptoms of depression, anxiety and stress. [21] It measures each item on a 4-point Likert scale. Items related to low positive affect, decreased self-esteem, hopelessness, and poor motivation are categorised as depression; while items related to autonomic arousal and fearfulness were categorised as anxiety. Items relating to irritability and over-reactivity are categorised as stress. [22] It has been validated in clinical and non-clinical populations. [23]

The Big Five Inventory (BFI-10) is a 10-item schedule to measure personality on the traits. [24] This is a 10-item scale with questions related to the five personality traits, answered on a 5 point Likert scale from “disagree strongly”, through “disagree a little”, “neither agree nor disagree”, “agree a little” to “agree strongly”. It is an abbreviated version of the Big Five Inventory (John et al, 1991). [25]

Data were analysed using the Statistical Package for the Social Sciences (SPSS) (Version 27.0). To make comparison between groups we examined basic bivariate characteristics. Stepwise linear regression analysis was used to investigate the association of demographic variables, personality and psychiatric morbidity with burnout after controlling for the population characteristics. To investigate these hypotheses we performed further multivariable linear regression analysis with the DASS outcomes (depression, anxiety and stress) as predictors and the three burnout dimensions as the dependent variable.

## Results

The characteristics of this group were previously described. [19] Of the 2,160 consultants invited to participate from the database of the Irish Hospital Consultants Association, 472 commenced the survey (response rate = 21.9%), and 433 completed the questionnaire: these were suitable for analysis. The socio-demographic characteristics of the group are outlined in Table 1. Of the respondents, 56.6% were male and 43.3% were female doctors, the mean age was 49.1 years and the majority were married or in a long-term relationship (82.7%). Medical physicians were the most represented specialty (19.4%), followed by psychiatrists (18.7%) and surgeons (15.7%). Just under one-third of respondents reported having been involved in a work-related legal action (31.9%), and 2.1% reported an experience of formal disciplinary action in the workplace.

**Table 1:** Demographics and work related characteristics

| **Characteristics** |  |  |
| --- | --- | --- |
| **Personal characteristics** |  |  |
| Age (years), mean (SD) |  | 49.14 (6.9) |
| Gender, n (%) | Male<br>Female | 253 (56.6)<br>194 (43.4) |
| Region, n (%) | Dublin<br>Munster<br>Connacht<br>Leinster<br>Ulster | 177 (39.2)<br>139 (30.6)<br>63 (14.0)<br>57 (12.6)<br>15 (3.3) |
| Relationship status, n (%) | Married<br>Long term relationship<br>Single<br>Divorced/separated<br>Widowed | 374 (82.7)<br>16 (3.5)<br>33 (7.3)<br>25 (5.5)<br>4 (0.9) |
| Habits, n (%) | Smoking<br>Alcohol<br>Exercise | 17 (3.7)<br>389 (87.8)<br>355 (78.9) |
| **Work characteristics** |  |  |
| Specialty, n (%) | Medicine<br>Psychiatry<br>Surgery<br>Anaesthesia<br>Radiology<br>Paediatrics<br>Pathology<br>Obstetrics & Gynaecology<br>Emergency Medicine<br>Ophthalmology | 86 (19.9)<br>81 (18.7)<br>68 (15.7)<br>47 (10.9)<br>46 (10.6)<br>30 (6.9)<br>29 (6.7)<br>25 (5.8)<br>16 (3.7)<br>5 (1.2) |
| Hours spent in "face-to-face" with patients per day, mean (SD) |  | 5.5 (4.3) |
| Public or private practice, n (%) | Wholly in the public system<br>Part public/ private<br>Part academic/ public/ private<br>Solely private<br>Part academic/ part public<br>Retired<br>Locum | 206 (46.5)<br>173 (39.1)<br>28 (6.3)<br>18 (4.1)<br>12 (2.7)<br>4 (0.9)<br>2 (0.5) |
| Satisfaction with remuneration, n (%) |  | 209 (47.6) |
| Formal disciplinary action at work, n (%) |  | 9 (2.1) |
| Work-related lawsuit, n (%) |  | 140 (31.9) |
| Hours worked/week, mean (SD) | Total<br>Public setting<br>Private<br>Managerial<br>Academic<br>Other | 51.8 (11.5)<br>30.9 (18.2)<br>7.25 (11.4)<br>3.76 (6.5)<br>2.1 (4.6)<br>0.83 (3.7) |

Although mean DASS scores for the whole group were below the cut off for caseness, 25.8% of hospital consultants reported levels of depressive symptoms above the cut-off and 13.8% reported anxiety symptoms above the cut-off. Almost 60% of participants reported stress levels above the cut off for caseness (See Table 2). The mean BFI (personality) scores are also shown in Table 2 and are significantly higher than in a normative general population sample where means sit between 3.2 (SD 0.83) for agreeableness and 4.1 (SD 0.69) for conscientiousness. [26] In this study population these means were 7.4 (SD 1.7) and 9.0 (1.3) respectively, more than double the general population scores in the literature.

**Table 2:** Mean scores on scales measuring depression, anxiety and stress (DASS) and personality traits (BFI)

| Scale/ Subscale | Mean (SD) | N (%) above cut-off |
| --- | --- | --- |
| DASS total | 10.9 (9.7) |  |
| DASS depression subscale | 3.3 (4.0) | 107 (25.8%) |
| DASS anxiety subscale | 1.6 (2.5) | 58 (13.8%) |
| DASS stress subscale | 6.0 (4.4) | 245 (59.2%) |
| BFI - extraversion | 6.3 (2.1) |  |
| - agreeableness | 7.4 (1.7) |  |
| - conscientiousness | 9.0 (1.3) |  |
| - Neuroticism | 5.0 (2.2) |  |
| - Openness | 7.4 (1.5) |  |
| DASS = Depressive Anxiety Stress Scale<br>BFI – Big Five Inventory |  |  |

We performed linear regression to examine the association between the three dimensions of burnout, psychiatric morbidity, and its contributory factors. After controlling for age and gender, higher scores on the emotional exhaustion burnout subscale were significantly associated with elevated symptoms of depression on the DASS depressive symptom subscale (p< 0.001). These were also associated with higher rates of face-to-face patient contact (p =0.001). Individual doctors with reduced satisfaction with remuneration (p =0.001) were more likely to develop psychiatric morbidity (table 3).

**Table 3:** Linear regression models with Emotional exhaustion, Cynicism and Personal Efficacy as the dependent variables

|  | B | p-value | Confidence Interval |
| --- | --- | --- | --- |
| *Emotional exhaustion* |  |  |  |
| Age | -0.046 | 0.313 | -0.136 – 0.044 |
| Gender | 0.946 | 0.136 | -0.3 – 2.192 |
| **DASS depression** | **1.032** | **<0.001** | **0.882 – 1.182** |
| **Face-to-face** | **0.219** | **0.001** | **0.085 – 0.352** |
| **Satisfaction with remuneration** | **2.111** | **0.001** | **0.889 – 3.333** |
| *Cynicism* |  |  |  |
| Age | 0.014 | 0.746 | 2.799 – 13.922 |
| Gender | 0.742 | 0.227 | -0.073 – 0.102 |
| **DASS stress** | **1.033** | **<0.001** | **0.9 – 1.167** |
| **Face-to-face** | **0.152** | **0.022** | **0.022 – 0.281** |
| **Satisfaction with remuneration** | **2.681** | **<0.001** | **1.501 – 3.861** |
| *Professional Efficacy* |  |  |  |
| Age | -0.047 | 0.342 | -0.143 – 0.05 |
| Gender | -0.992 | 0.14 | -2.312 – 0.328 |
| **DASS stress** | **-0.459** | **<0.001** | **-0.607 – -0.311** |
| **Face-to-face** | **0.196** | **0.024** | **0.026 – 0.367** |
| Satisfaction with remuneration | -0.948 | 0.15 | -2.241 – 0.345 |

Elevated burnout scores on the cynical subscale were associated with high DASS stress symptoms (p< 0.001) after controlling for age and gender. Greater weekly hours of face-to-face patient contact was associated with higher scores on the cynicism subscale (p= 0.022) and poor satisfaction with remuneration was likewise associated with higher scores on the cynicism subscale (p< 0.001) as shown in table 3.

Professional efficacy is the third subscale of the MBI-GS and is a protective factor for the development of burnout. In this study we found that after controlling for age and gender there was an inverse relationship with the stress subscale of the DASS (p < 0.001), i.e., among those reporting higher professional efficacy, higher levels of stress were found. Professional efficacy is associated with lower levels of burnout. It was also significantly associated with elevated face-to-face time with patients (0.024), and there was no association in this model with satisfaction with remuneration and professional efficacy (table 3).

## Discussion

This study reported a high prevalence of burnout among hospital consultants in Ireland. High proportions screened positive for stress (60%), symptoms of depression (26%) and symptoms of anxiety (14%). Burnout is associated with higher scores of depressive symptoms and stress as measured by DASS. Higher levels of patient contact were associated with increased emotional exhaustion and cynicism, but also with elevated levels of the protective professional efficacy. Low levels of satisfaction with remuneration were associated with increased emotional exhaustion and cynicism, but there was no association with professional efficacy.

Using the composite measure of burnout, higher levels of burnout were found in this sample compared with those reported by O’Dea in a previous study of Irish general practitioners (GPs) (10.8% vs 6.6%). [20] O’Dea’s study reported higher levels of emotional exhaustion (52.7% compared with 42.7%) and lower levels of the professional efficacy which is protective for burnout (16.3% compared with 1.2%). [20] O’Dea et al found similar rates of burnout to UK-based GPs according to a European study, where UK GPs scored higher than those based in Greece, Turkey, Italy and Bulgaria. [27]

Compared to interns, Irish consultants appear have lower rates of burnout than more junior hospital doctors. Two-fifths 42% of Irish interns reported a low sense of professional efficacy, and a majority (51.5%) reported high rates of emotional exhaustion [24]. It is unknown whether the working conditions of younger doctors, or more general factors associated with age predispose them to burn-out. For example, there may be an association between age and working hours as junior doctors face greater pressure to increase knowledge and skills through practice. Furthermore, their lack of experience may mean they take longer to complete tasks, thereby resulting in increased working hours. [28]

The response rate in this study compares favourably with previous response rates in similar studies: Al Humadi et al recruited 225 senior doctors, a response rate of 16.3%, and Garcia et al reported a response rate of 17.1% in their US based studies [29, 30].

Al Humadi et al reported rates of 6.2% depression, 6.6% suicidal ideation, and 19.6% burnout during the Covid-19 pandemic, all associated with history of prior depression or anxiety. Suicidal ideation and burnout were also associated with younger age. Female physicians reported reduced work-life balance and higher rates of burnout. In our study gender was not found to be significantly associated with burnout. A more recent study of Irish consultants conducted during the Covid-19 pandemic in 2021, found that three-quarters screened positive for burnout in the context of the pandemic. [31]

The association of burnout with longer time spent face-to-face with patient may be a proxy for longer hours of clinical work (given that the mean amount of patient contact was 5.5 hours per day), or it may perhaps represent the higher emotional toll of patient care, especially in certain specialities. Previous studies have found a similar association between high clinical workload and burnout. [32]

High rates of work-related burnout in consultants are highly concerning. Symptoms of stress, anxiety and depression indicate the high psychological burden of burnout. Personality factors, particularly conscientiousness and agreeableness, appear to have an influence on physician burnout. There is a need for research to identify appropriate strategies (both short-term and long-term) to minimise the impact of burnout and to ensure physician wellbeing. There is an urgent need for evidence-based strategies perhaps modulating the relationship between personality and burnout, to mitigate the impact on symptoms of depression and anxiety and to optimise doctors’ health for optimal health care delivery.

Any interventions must include a consideration of both organisational and individual level measures. West et al conducted a systematic review of the evidence for the management of burnout and found strong evidence base for the use of mindfulness, stress management programmes and small group (including reflective) discussions [33]. There was limited evidence for organisational interventions, mainly due to a lack of studies: it is much more challenging to perform a randomised controlled trial of organisational change than a mindfulness programme [33, 34]. At an individual level, doctors tend to identify organisational and structural factors as the priority areas for improving their working lives and addressing burnout [35].

The limitations of this study include its cross-sectional design and the use of DASS, a screening rather than a diagnostic measure. The small response rate, while not dissimilar to other similar studies, suggests the potential for response bias: people feeling the effects of burnout may be more likely to respond to a survey on this topic, but conversely doctors who are feeling especially overstretched may not feel able to take on another task. It is, however, a large national sample of senior clinicians which includes broad representation across geographical location and speciality.

Given these high rates of burnout and stress that were reported pre-covid, it would be important to consider the changes that have occurred in healthcare since 2020. There is an urgent need to consider the need to consider measures to mitigate the impact of burnout and to consider the causes, especially structural causes that it may be possible to mitigate.

Burnout is common in senior doctors, and it is associated with high levels of stress and symptoms of depression and anxiety. Given that physician mental health and suicide is an internationally growing area of concern, these findings suggest that burnout is associated with poorer mental health. There is an urgent need for more research on the relationship between burnout and depression and anxiety in the post-covid world and for the introduction of measures to mitigate its effect on doctors and patients.

## Key points

### What is already known about this subject

- Burnout is a common problem among healthcare professionals especially doctors and includes three domains: emotional exhaustion, cynicism and professional efficacy.
- Burnout is associated with poorer mental and physical health, and people who meet the threshold for burnout frequently endorse symptoms of depression.

### What this study adds

- High-levels of burnout and stress were reported in this study of senior doctors (consultants) in this pre-covid study.
- Emotional exhaustion is associated with depressive symptoms after controlling for age, gender and higher workload.

### What impact this may have on practice or policy

- Addressing factors which contribute to burnout may have a positive impact on physician mental health.
- Work-related factors must be considered in any strategies designed to address physician burnout.

## Data Availability

Data cannot be share publicly because given the level of data (including job characteristics i.e. there is only one consultant liaison psychiatrist in Galway), participants are potentially identifiable from the research dataset. Data are available on request to the corresponding author.

## Author contributions

Conceptualisation: GC, FM AMD. Data curation: GC, FM AMD. Formal analysis: GC, AMD. Investigation: GC, FM AMD. Methodology: GC, FM AMD. Project administration: GC, FM. Supervision: AMD. Visualisation : GC, FM AMD. Writing - original draft: GC. Writing – review and editing: GC, FM, AMD.

## Acknowledgements

The authors acknowledge the Irish Hospital Consultants Association for allowing access to the membership email, and to its members for participating in this study.

## Competing interests

The authors have no interests to declare.

## Funding

This research did not receive any specific grant from funding agencies in the public, commercial, or not-for-profit sectors.

## Notes

### Competing Interest Statement

The authors have declared no competing interest.

### Funding Statement

The author(s) received no specific funding for this work.

### Author Declarations

The project received ethical approval from the Saolta Research Ethics Committee (CA1591). The study was conducted in line with the principles of the Declaration of Helsinki.

